# Accelerated Epigenetic Aging is Associated with Faster Glaucoma Progression: A DNA Methylation Study

**DOI:** 10.1101/2024.10.12.24315371

**Authors:** Felipe A. Medeiros, Achintya Varma, Alessandro A. Jammal, Henry Tseng, William K. Scott

## Abstract

**Purpose:** To investigate the association between epigenetic age acceleration and glaucoma progression.

**Design:** Retrospective cohort study.

**Participants:** 100 primary open-angle glaucoma (POAG) patients with fast progression and 100 POAG patients with slow progression.

**Methods:** Subjects were classified as fast or slow progressors based on rates of change in standard automated perimetry (SAP) mean deviation (MD) and retinal nerve fiber layer (RNFL) thickness. Epigenetic age was calculated using the Horvath, Hannum, PhenoAge, and GrimAge clocks from DNA methylation profiles obtained from blood samples. Age acceleration (AgeAccel) was defined as the residual from a linear regression of epigenetic age on chronologic age, with positive values suggesting faster biological aging. Multivariable logistic regression models estimated the association between AgeAccel and likelihood of fast progression, adjusting for confounders.

**Main Outcome Measures:** Difference in epigenetic age acceleration between fast and slow glaucoma progressors.

**Results:** The mean rate of SAP MD change in the fastest progressing eye was −1.06 dB/year (95% CI: −1.28 to −0.85) for fast progressors compared to −0.10 dB/year (95% CI: −0.16 to −0.04) for slow progressors (P<0.001). For RNFL thickness, corresponding values were −1.60 μm/year (95% CI: −1.97 to −1.23) and −0.76 μm/year (95% CI: −1.04 to −0.48), respectively (P<0.001). Fast progressors demonstrated significantly greater age acceleration compared to slow progressors for the Horvath clock (mean difference = 2.93 years, 95% CI: 1.48 to 4.39, P<0.001) and Hannum clock (mean difference = 1.24 years, 95% CI: 0.03 to 2.46, P=0.045). In multivariable models, each year of Horvath AgeAccel was associated with 15% higher odds of fast progression (OR 1.15, 95% CI 1.07-1.23, P<0.001), after adjusting for sex, race, intraocular pressure, central corneal thickness, baseline disease severity, smoking status and follow-up time. Hannum and GrimAge clocks also showed significant associations with fast progression. The association between AgeAccel and fast progression was stronger in subjects with relatively low IOP during follow-up.

**Conclusion:** Accelerated epigenetic aging was associated with faster glaucoma progression. These findings suggest that faster biological age, as reflected in DNA methylation, may increase optic nerve susceptibility to damage, highlighting epigenetic age as a potential prognostic biomarker.

## INTRODUCTION

Glaucoma is a neurodegenerative disease characterized by progressive loss of retinal ganglion cells and their axons.^1^ The disease affects over 100 million individuals worldwide and is the leading cause of irreversible blindness. Although elevated intraocular pressure (IOP) is the most widely accepted modifiable risk factor for glaucoma, a significant proportion of patients continue to experience glaucomatous deterioration despite well-controlled IOP, suggesting that other factors contribute to the progression of glaucomatous damage.^2^ Moreover, a subset of 30% to 40% of patients exhibits normal-tension glaucoma, where damage occurs without concomitant apparent IOP elevation. This indicates that factors other than IOP elevation may increase the optic nerve’s susceptibility to damage.^3^

Aging plays a pivotal role in glaucoma. The prevalence of glaucoma increases markedly with age.^4, 5^ However, the pathophysiology underlying increasing age as a risk factor for glaucoma is not well understood. The increased prevalence of glaucoma in older individuals does not seem to be explained solely by an increased prevalence of high IOP with aging.^6–8^ This suggests that aging may increase the vulnerability of the optic nerve to IOP-related damage, ultimately resulting in loss of retinal ganglion cells. This age-related increased vulnerability to neural injury has also been observed in other neurodegenerative disorders, such as Alzheimer’s and Parkinson’s disease, and may be related to mitochondrial dysfunction and impaired capacity to handle oxidative stress, among other factors.^9–15^ Epigenetics, the study of alterations in gene expression without changes in the DNA sequence, has shown promise in understanding aging and age-related diseases. The biological age encapsulated in our epigenome, as reflected by DNA methylation patterns, often diverges from our chronological age. The “epigenetic clock” theory, conceptualized by Horvath,^16^ leverages these methylation changes at specific cytosine-phosphate-guanine (CpG) sites as a timekeeper of biological aging. The disparities between biological (epigenetic) and chronological age have been associated with several age-related diseases, such as cardiovascular disease, neurodegeneration, and cancer. An advanced epigenetic age relative to chronological age may signify accelerated biological aging and increased disease risk.^17^ Despite these observations, no study has yet been conducted on epigenetic age and glaucoma.

In the present study, we hypothesized that disparities between epigenetic and chronological age may be a significant risk factor in explaining glaucoma progression. Specifically, we hypothesized that individuals with faster disease progression would demonstrate an accelerated biological age, as indicated by a greater divergence between epigenetic and chronological age, compared to those with slower progression. Uncovering the relationship between epigenetic age and glaucoma progression could provide novel insights into the pathophysiology of the disease and offer a more accurate prognostic tool than chronological age alone.

## METHODS

This study involved a cohort of subjects that were part of the Duke Glaucoma Registry (DGR), a large database of electronic medical records developed at Duke University, Durham, North Carolina. The database consisted of adults 18 years or older with glaucoma or glaucoma suspect diagnoses who were evaluated at the Duke Eye Center or its satellite clinics between January 2009 and June 2023. The database has been described in detail elsewhere and has been used for several studies investigating risk factors for glaucoma.^3, 18–21^ A subset of participants, described below, was recruited for blood sample collection. Blood sample analyses were performed at the John P. Hussman Institute for Human Genomics (HIHG) at the University of Miami, Florida. The Duke University and the University of Miami Institutional Review Boards approved this study. Informed consent was obtained from all participants. All methods adhered to the tenets of the Declaration of Helsinki for research involving human subjects and were conducted in accordance with regulations of the Health Insurance Portability and Accountability Act.

The database used for this study contained clinical information from baseline and follow-up visits, including patient diagnostic and procedure codes, medical history, smoking history, best-corrected visual acuity, slit-lamp biomicroscopy, IOP measurement using the Goldmann applanation tonometry (GAT; Haag-Streit, Konig, Switzerland), central corneal thickness (CCT), gonioscopy, ophthalmoscopy examination, optic disc photographs, and the results of all standard automated perimetry (SAP) and optical coherence tomography (OCT) exams. SAP testing was performed with the Humphrey Field Analyzer (Carl Zeiss Meditec, Dublin, CA) using the Swedish Interactive Thresholding Algorithm (SITA) 24-2 fast strategy. OCT was performed with the Spectralis SDOCT (Heidelberg Engineering, Dossenheim, Germany). A reliable SAP was defined as having fixation loss rate less than 25% and false-positive rate less than 15%. Visual fields were manually reviewed for artifacts such as lid and rim artifacts, fatigue effects, inappropriate fixation, and evidence that the visual field results were due to a disease other than glaucoma. Retinal nerve fiber layer (RNFL) thickness measurements were obtained from a 12-degree (for single circle scans) or a 3.45mm-diameter peripapillary circle scan (for scans from the Glaucoma Mode Premium Edition) acquired using the Spectralis SD OCT, as described in detail previously.^22^ Tests were acquired using the latest available software version at the time of the scan and exported using the latest available version at the time of the analysis (Software version 6.8). Scans were manually reviewed for the presence or artifacts, decentration, or segmentation errors. Tests with quality score below 15, or with segmentation errors, evidence of decentration or artifacts were also excluded. For each scan, the global average RNFL thickness was calculated as the average of thicknesses of all points from the 360 degrees around the optic nerve head.

### Participant selection

This study included a subset of participants from the DGR who were recruited for prospective blood sample collection. Subjects were contacted during their regular clinic visits and invited to provide blood samples for genetic analysis. Selection criteria included a diagnosis of primary open-angle glaucoma (POAG) in both eyes, confirmed by international classification of diseases (ICD) codes at the baseline visit, as well as the availability of longitudinal data in the DGR. 1,251 POAG patients provided blood samples. A total of 10 ml of blood was collected from each participant, processed, and stored for future analysis. Blood collections took place between June 2021 and June 2023.

### Fast versus slow progression classification

Subjects were classified as either fast or slow progressors based on available SAP and SD OCT tests. Ordinary least squares linear regression was used to estimate the slopes of change in SAP mean deviation (MD) and RNFL thickness over time. Subjects who did not clearly fit into either group were excluded from further analyses to maximize the distinction between fast and slow progressors and avoid including ambiguous cases. Fast progressors were defined as having at least one eye with either a statistically significant SAP MD slope faster than −0.5 dB/year or a statistically significant global RNFL thickness slope faster than −1.0 µm/year. In contrast, slow progressors were required to meet all of the following criteria in both eyes: non-significant SAP MD slope, non-significant global RNFL thickness slope, a minimum follow-up time of 5 years, and no history of trabeculectomy or tube shunt surgery.

These criteria were designed to create a clearly divergent sample, thereby enhancing the ability to test the primary hypothesis of the study. No minimum follow-up time was required for fast progressors, allowing the inclusion of patients with rapid progression over shorter periods. While a history of glaucoma surgery was not an exclusion criterion for fast progressors, SAP and OCT tests were censored post-surgery and excluded from the calculation of progression rates. Importantly, we use the term ‘slow progressor’ rather than ‘non-progressor’ because it is not possible to guarantee that these subjects would not have progressed by other measures. The primary objective was to ensure that any changes in the slow progressor group would likely be minimal, resulting in a significant difference in disease trajectory between the two groups.

### Sample Size Calculation

The primary hypothesis of this study was that individuals with faster glaucoma progression would demonstrate an accelerated biological age, indicated by a greater discrepancy between epigenetic age and chronological age, compared to those with slower progression. Given the high costs associated with DNA methylation analysis, only a subset of subjects with available blood samples underwent epigenetic age calculation. A power analysis determined that a sample size of 100 subjects per group would provide 80% power to detect a minimum difference of 2.0 years between the two groups in the difference between epigenetic and chronological age, assuming a standard deviation of 5.0 and an alpha level of 0.05. Consequently, from the 1,251 subjects with available blood samples, 100 individuals with slow progression and 100 with fast progression were selected. These 200 samples were then subjected to DNA extraction, methylation profiling and epigenetic age calculation.

### DNA Extraction, Methylation Processing, and Quality Control

Immediately after collection, whole blood samples were centrifuged to separate the different components. The separated components, including plasma, buffy coat, and red blood cells, were then frozen at −80°C for preservation. At a later stage, the buffy coat was thawed and processed for DNA extraction. Upon thawing, the buffy coat was centrifuged at 1,500 x g for 10 minutes to concentrate the cells. Genomic DNA was subsequently extracted from the buffy coat using the QIAamp DNA Blood Mini Kit (Qiagen, Hilden, Germany), following the manufacturer’s protocol. The quality of the extracted DNA was assessed using a NanoDrop spectrophotometer (Thermo Fisher Scientific, Waltham, MA). The absorbance ratios at 260/280 nm and 260/230 nm were measured, with DNA samples considered high quality if the A260/280 ratio fell between 1.8 and 2.0, and the A260/230 ratio was ≥2.0. The quantity and concentration of DNA were evaluated using a Qubit Fluorometer (Thermo Fisher Scientific, Waltham, MA). Additionally, DNA integrity was qualitatively evaluated using agarose gel electrophoresis to identify samples with high molecular weight bands suitable for analysis. Sample concentration was normalized to 50 ng/μL and arrayed in Azenta 0.5 ml barcoded tubes in racks of 96.

Genome-wide DNA methylation was assessed using the Illumina Infinium MethylationEPIC v2 BeadChip (Illumina, San Diego, CA), which interrogates approximately 930,000 cytosine-phosphate-guanine (CpG) sites across the human genome. DNA samples (500 ng each) were bisulfite-converted using the EZ DNA Methylation Kit (Zymo Research, Irvine, CA) according to the manufacturer’s instructions, which converts unmethylated cytosines to uracil while leaving methylated cytosines unchanged. The bisulfite-converted DNA was then hybridized to the MethylationEPIC v2 BeadChip, and the arrays were scanned using the Illumina iScan system. Data files created by the Illumina iScan were analyzed using the Illumina GenomeStudio Methylation module software program. To ensure the reliability of the methylation data, several quality control measures were implemented following established protocols in the literature. Raw methylation intensity data were first preprocessed using the R package minfi, which includes background correction, normalization, and calculation of detection p-values to assess the quality of individual probes.^23^ Probes with detection p-values above a threshold of 0.01 in more than 5% of samples were excluded from further analysis.^24^ Methylation data were then mapped to the genome, and only those probes that passed stringent quality control criteria were retained for epigenetic age calculation.

### Epigenetic Age Calculation

Epigenetic age was calculated using several established algorithms, each of which applies specific sets of CpG sites to estimate biological age:

- **Horvath Clock**: The Horvath clock is an epigenetic age estimator that utilizes a linear combination of methylation levels at 353 specific CpG sites to calculate an individual’s biological age; these sites were selected from data generated from multiple tissue types, including blood, skin, brain and other internal organs.^25^ This “universal” epigenetic clock is designed to provide a robust estimate of biological age, regardless of tissue source, and has been widely validated for its accuracy in correlating with chronological age across a diverse range of tissues.
- **Hannum Clock**: The Hannum clock is an epigenetic age predictor specifically tailored for use in whole blood samples.^26^ It is based on methylation data from 71 CpG sites that are strongly associated with age-related changes in blood. The model calculates biological age by assessing methylation patterns at these sites, which reflect the physiological aging process in hematopoietic cells, making it potentially useful for studies focusing on blood-based biomarkers of aging.
- **PhenoAge**: The PhenoAge clock, developed by Levine et al.,^27^ integrates DNA methylation data from 513 CpG sites with clinical phenotypes and mortality risk indicators to provide a comprehensive measure of biological aging. Unlike traditional epigenetic clocks that primarily estimate chronological age, PhenoAge is designed to capture the physiological decline associated with morbidity and mortality risk.
- **GrimAge**: GrimAge is an epigenetic clock that combines methylation data from specific CpG sites with inferred plasma protein levels and cumulative smoking exposure to predict time to death and other aging-related outcomes.^28^

Methylation data for the probes was submitted to the DNA Methylation Age Calculator website (dnamage.clockfoundation.org).^25^ For each one of these clocks, age acceleration (AgeAccel) was calculated as the residual from a linear regression model of the corresponding epigenetic clock and chronological age. Residual variables were then named AgeAccelHorvath, AgeAccelHannum, AgeAccelPheno and AgeAccelGrim, corresponding to the clocks listed above. Positive values in these variables suggest a higher epigenetic age relative to chronological age and thus faster biological aging, while negative values suggest slower biological aging.

### Statistical Analysis

Correlations between chronological age and each epigenetic age measure were assessed using Pearson’s correlation coefficients. Differences in AgeAccel (epigenetic age acceleration) between fast and slow glaucoma progressors were evaluated using Student’s t-tests. Additionally, multivariable logistic regression models were employed to estimate adjusted odds ratios (ORs) and 95% confidence intervals (CIs) for the association between AgeAccel variables and the likelihood of fast progression. These models were adjusted for key potential confounders, including gender, race, smoking status, IOP, CCT, and baseline disease severity, as measured by baseline MD. For IOP, both the mean and peak values recorded during follow-up were analyzed. Since IOP, CCT, and baseline MD are eye-specific variables, the mean values of both eyes were utilized in the logistic regression models. Furthermore, considering that blood samples were collected during follow-up rather than at baseline, the time from baseline to the date of blood collection was calculated and included as a potential confounder in the multivariable models.

Statistical analyses were completed in R (version 4.4.1, R Foundation for Statistical Computing, Vienna, Austria).

## RESULTS

The study included 100 POAG patients diagnosed with fast glaucoma progression and 100 POAG patients with slow progression. **Table 1** shows demographic and clinical characteristics of the 2 groups. Mean follow-up time was 10.3 years (95% CI: 9.0 – 11.5) for fast progressors and 11.0 years (95% CI: 9.6 – 12.3) for slow progressors, with no statistically significant difference between the groups (P = 0.45). The mean rate of change in SAP MD, averaged across both eyes, was significantly different between the groups: −0.63 dB/year (95% CI: −0.77 – −0.48) in fast progressors compared to 0.01 dB/year (95% CI: −0.06 – 0.07) in slow progressors (P < 0.001). When considering the eye with the faster rate of SAP MD decline, the mean rates were −1.06 dB/year (95% CI: −1.28 – −0.85) for fast progressors and −0.10 dB/year (95% CI: −0.16 – −0.04) for slow progressors (P < 0.001). Over the follow-up period, fast progressors lost an average of 8.2 dB (95% CI: 6.9 – 9.5) of MD in the fastest progressing eye, compared to a loss of only 1.4 dB (95% CI: 0.8 – 1.9) in the slow progressor group. For OCT RNFL thickness, the mean rate of change in the fastest progressing eye was −1.60 µm/year (95% CI: −1.97 – −1.23) for fast progressors, compared to −0.76 (95% CI: −1.04 – −0.48) µm/year for slow progressors (P < 0.001). Mean IOP during follow-up was similar between the groups, at 15.0 mmHg (95% CI: 14.4 – 15.6) for fast progressors and 15.3 mmHg (95% CI: 14.7 – 16.0) for slow progressors (P = 0.39). However, peak IOP during follow-up, averaged across both eyes, was higher in fast progressors compared to slow progressors (23.8 mmHg [95% CI: 22.0 – 25.5) vs. 20.5 mmHg [95% CI: 19.5 – 21.6], respectively; P = 0.002).

**Table 1.** Demographic and clinical characteristics of fast and slow glaucoma progressors.

| Characteristic | Slow Progressors<br>(n=100) | Fast Progressors<br>(n=100) | P |
| --- | --- | --- | --- |
| Age at baseline (years) | 61.8 ± 9.0 | 60.6 ± 11.8 | 0.43 |
| Age at blood collection (years) | 73.4 ± 8.3 | 71.7 ± 10.2 | 0.20 |
| Sex (female), n (%) | 59 (59%) | 58 (58%) | 0.89 |
| Race (Black or African American), n (%) | 21 (21%) | 22 (22%) | 0.86 |
| Smoking status, yes, n (%) | 5 (5%) | 2 (2%) | 0.25 |
| Baseline MD (dB) | -3.52 ± 4.52 | -6.05 ± 5.75 | <0.001 |
| Peak IOP (mmHg) | 20.5 ± 5.4 | 23.8 ± 8.6 | 0.002 |
| Mean IOP (mmHg) | 15.3 ± 3.3 | 15.0 ± 3.1 | 0.39 |
| CCT (µm) | 548.7 ± 34.0 | 542.5 ± 45.9 | 0.28 |
| Number of tests | 8.1 ± 5.0 | 8.3 ± 4.4 | 0.81 |
| Follow-up time (years) | 11.0 ± 6.7 | 10.3 ± 6.1 | 0.45 |
| Time to blood collection (years) | 11.5 ± 6.6 | 11.4 ± 6.3 | 0.86 |
| Average MD slope (dB/year) | 0.01 ± 0.32 | -0.63 ± 0.72 | <0.001 |
| Fastest MD slope (dB/year) | -0.10 ± 0.33 | -1.06 ± 1.08 | <0.001 |
| Average RNFL slope (µm/year) | -0.30 ± 1.42 | -0.99 ± 1.59 | 0.002 |
| Fastest RNFL slope (µm/year) | -0.76 ± 1.37 | -1.60 ± 1.86 | <0.001 |

Values indicate mean ± standard deviation unless otherwise noted. CCT: central corneal thickness, IOP: intraocular pressure, MD: mean deviation, RNFL: retinal nerve fiber layer thickness.

Epigenetic age was strongly correlated with chronological age across all epigenetic clock methods (**Figure 1**). The Horvath clock showed a Pearson correlation coefficient of r = 0.83 (95% CI: 0.78 – 0.87) with chronological age (P<0.001). Similarly, the Hannum clock demonstrated a correlation of r = 0.85 (95% CI: 0.80 – 0.90; P<0.001), while the PhenoAge clock showed a correlation of r = 0.81 (95% CI: 0.76 – 0.87; P<0.001). The GrimAge clock had a slightly lower but still substantial correlation with chronological age (r = 0.70; 95% CI: 0.62 – 0.78; P<0.001).

**Figure 1.**
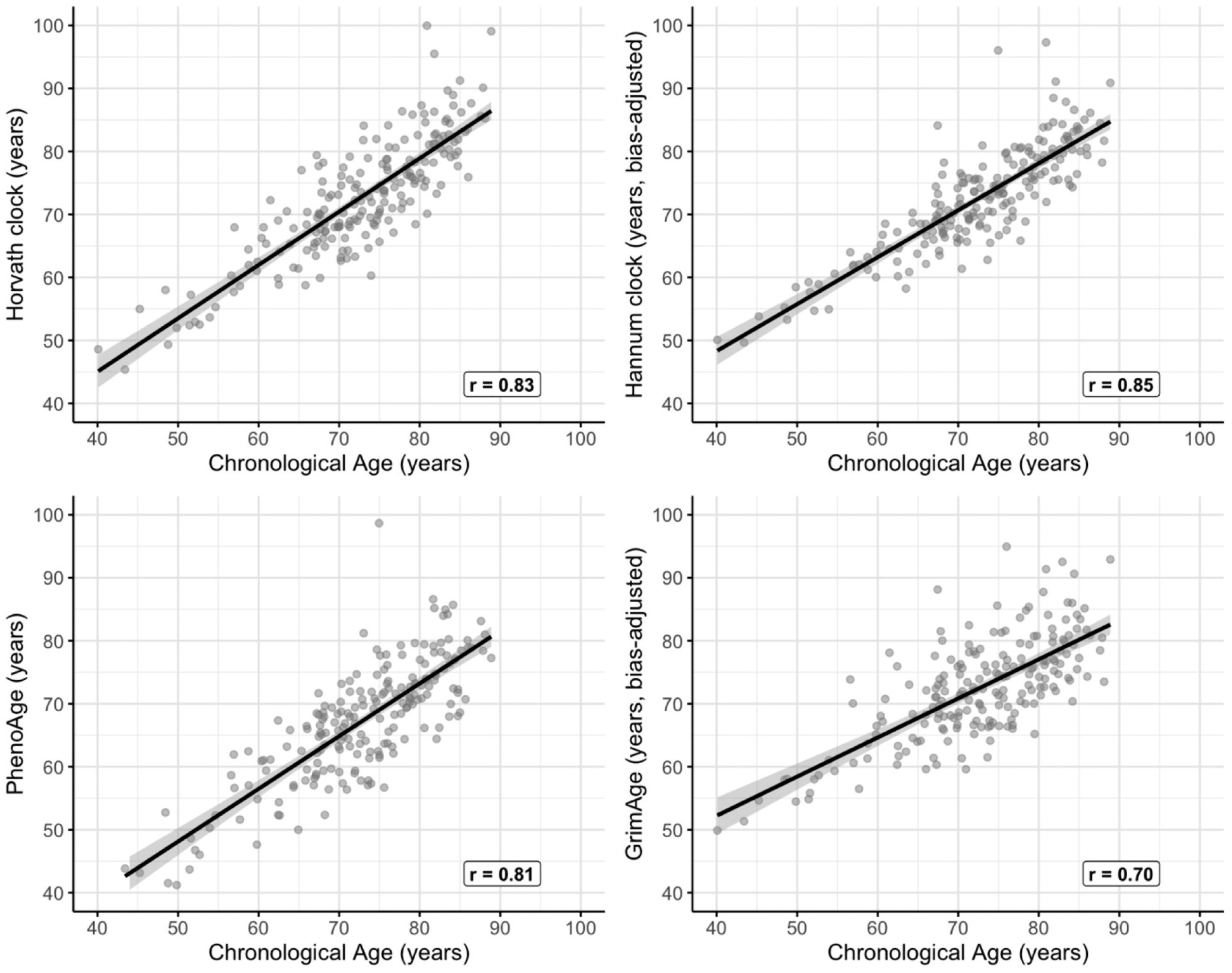
Scatter plots illustrating the relationship between epigenetic age (y-axis) and chronological age (x-axis) for four epigenetic clocks: (A) Horvath, (B) Hannum, (C) PhenoAge, and (D) GrimAge. Each point represents an individual patient. Pearson correlation coefficients (r) are shown for each clock. Epigenetic age was calculated using DNA methylation data from blood samples collected during the follow-up period. Chronological age is the age at blood collection.

**Table 2** shows the mean age acceleration values for fast and slow progressors according to the different epigenetic clocks, while **Figure 2** shows the distribution of these values across the two groups. For AgeAccelHorvath, fast progressors had significantly greater age acceleration compared to those with slow progression, with a mean difference of 2.93 years (95% CI: 1.48 to 4.39; P < 0.001). Similarly, faster acceleration was observed with AgeAccelHannum, with a mean difference of 1.24 years (95% CI: 0.03 to 2.46; P = 0.045). No statistically significant differences were found for AgeAccelPheno or AgeAccelGrim. For AgeAccelPheno, the mean difference between fast and slow progressors was 1.06 years (95% CI: −0.51 to 2.64; P = 0.185). Likewise, AgeAccelGrim showed a mean difference of 1.49 years (95% CI: −0.17 to 3.14; P = 0.079), which also did not reach statistical significance.

**Figure 2.**
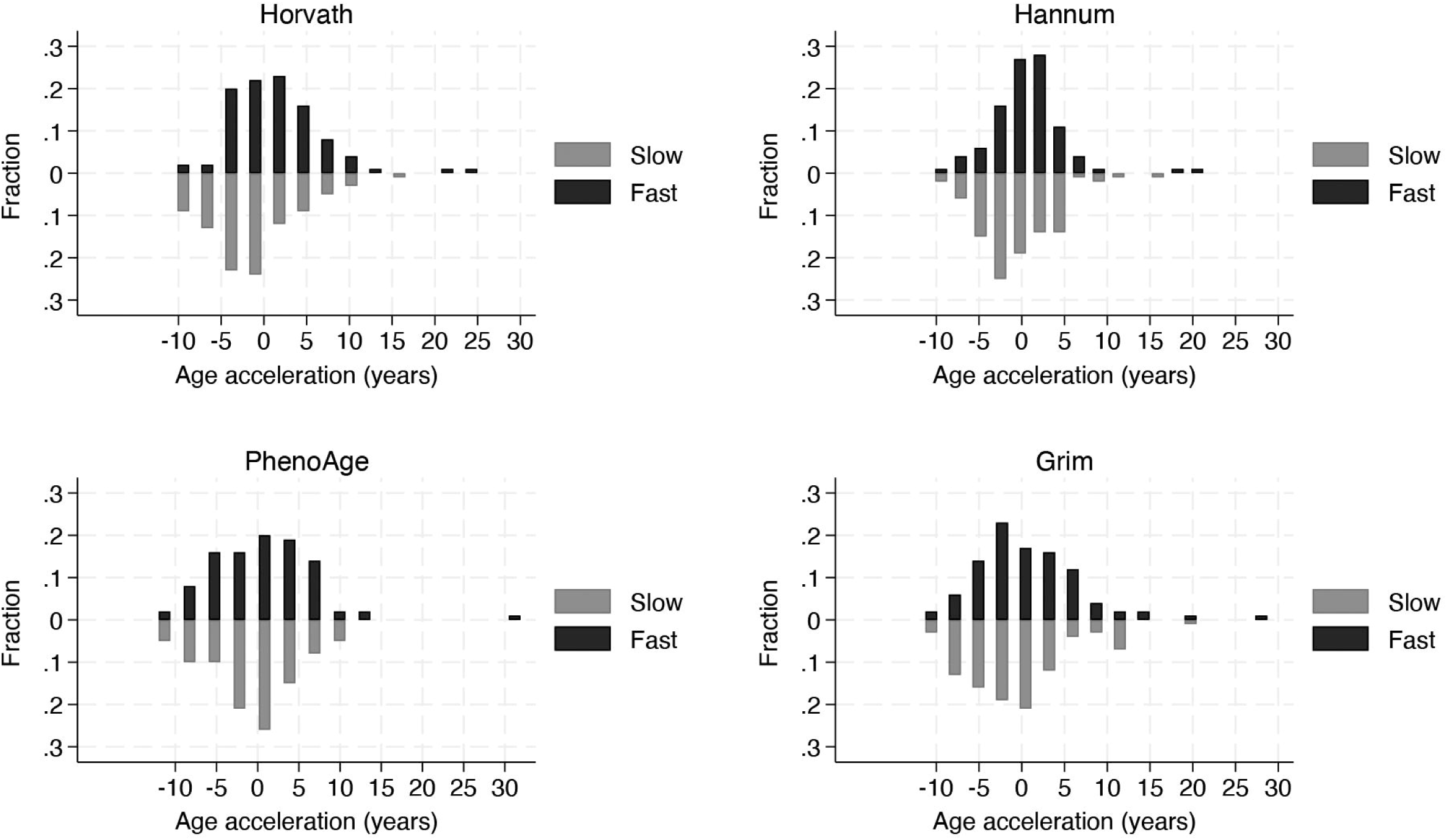
Histograms depicting the distribution of epigenetic age acceleration values for (A) Horvath, (B) Hannum, (C) PhenoAge, and (D) GrimAge clocks in fast (black) and slow (grey) glaucoma progressors. Age acceleration was calculated as the residual from a linear regression of epigenetic age on chronological age. Positive values indicate an epigenetic age greater than chronological age.

**Table 2.**
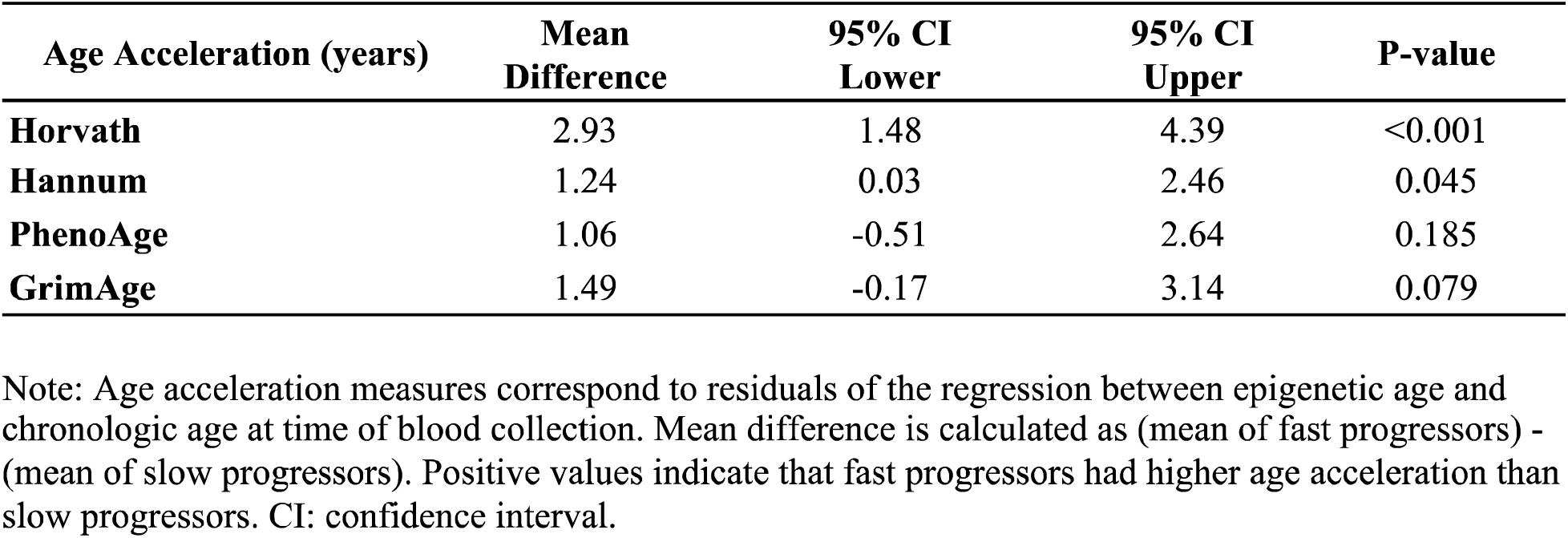
Differences in epigenetic age acceleration between fast and slow glaucoma progressors.

**Table 3** shows the results of multivariable logistic regression models for the association between different age acceleration variables and the likelihood of fast progression, adjusted for relevant covariates. AgeAccelHorvath demonstrated the strongest association with fast progression (OR: 1.15 per year of acceleration, 95% CI: 1.07-1.23, P<0.001). AgeAccelHanum and AgeAccelGrim also showed significant associations (OR: 1.07, 95% CI: 1.00-1.15, P=0.049 and OR: 1.07, 95% CI: 1.01-1.14, P=0.034, respectively). AgeAccelPheno did not reach statistical significance (OR: 1.04, 95% CI: 0.99-1.10, P=0.141). Lower baseline MD, indicating worse severity at baseline, was significantly associated with fast progression across all models (OR ranging from 1.12 to 1.13 per dB lower, P=0.001 for all models). Peak IOP was also a significant predictor in all models, with the OR consistently at 1.09 or 1.10 per mmHg increase (P≤0.001 for all models). Male sex showed a trend towards higher odds of fast progression, although this did not reach statistical significance. CCT, race, follow-up time, and smoking status were not significantly associated with fast progression in any of the models.

**Table 3.** Results of multivariable logistic regression models for the association between epigenetic age acceleration and fast glaucoma progression.

| Variable | Horvath |  | Hannum |  | PhenoAge |  | Grim |  |
| --- | --- | --- | --- | --- | --- | --- | --- | --- |
|  | OR (95% CI) | P | OR (95% CI) | P | OR (95% CI) | P | OR (95% CI) | P |
| Age Acceleration, y | 1.15 (1.07-1.23) | <0.001 | 1.07 (1.00-1.15) | 0.049 | 1.04 (0.99-1.10) | 0.141 | 1.07 (1.01-1.14) | 0.034 |
| Sex, male | 1.56 (0.79-3.07) | 0.197 | 1.47 (0.76-2.87) | 0.254 | 1.29 (0.68-2.45) | 0.436 | 1.59 (0.81-3.15) | 0.181 |
| Race, Black/AA | 1.03 (0.48-2.23) | 0.935 | 1.11 (0.52-2.34) | 0.789 | 0.92 (0.42-1.98) | 0.824 | 0.82 (0.37-1.79) | 0.617 |
| Peak IOP | 1.10 (1.04-1.15) | <0.001 | 1.09 (1.04-1.15) | 0.001 | 1.09 (1.04-1.15) | 0.001 | 1.09 (1.04-1.14) | 0.001 |
| Baseline MD, per dB lower | 1.13 (1.05-1.22) | 0.001 | 1.13 (1.05-1.21) | 0.001 | 1.13 (1.05-1.21) | 0.001 | 1.12 (1.05-1.20) | 0.001 |
| CCT, per 40µm thinner | 1.23 (0.88-1.70) | 0.223 | 1.19 (0.87-1.64) | 0.274 | 1.19 (0.87-1.63) | 0.282 | 1.26 (0.91-1.74) | 0.164 |
| Follow-up time, y | 1.03 (0.98-1.08) | 0.312 | 1.02 (0.97-1.07) | 0.471 | 1.02 (0.97-1.07) | 0.504 | 1.02 (0.97-1.07) | 0.380 |
| Smoking, yes | 0.46 (0.08-2.63) | 0.380 | 0.56 (0.10-3.17) | 0.512 | 0.58 (0.10-3.28) | 0.537 | 0.35 (0.06-2.14) | 0.255 |

AA: African-American, CCT: central corneal thickness, CI: confidence interval, IOP: intraocular pressure, MD: mean deviation, OR: odds ratio. Follow-up time: follow-up time until blood collection

We further examined the effect of age acceleration on the likelihood of fast progression in POAG subjects who had no history of elevated IOP, defined as those with both eyes never exceeding an IOP of 21 mmHg (**Table 4, supplemental**). Statistically significant differences were observed across all age acceleration metrics, except for AgeAccelPheno. Specifically, for AgeAccelHorvath, the mean difference between fast and slow progressors was 4.69 years (95% CI: 2.46 to 6.93), indicating acceleration in epigenetic aging among fast progressors.

## DISCUSSION

To our knowledge, this study is the first to explore the link between epigenetic age and glaucoma progression, offering new insights into the role of biological aging in this neurodegenerative disease. Our findings revealed a significant association between accelerated epigenetic age and faster glaucoma progression, suggesting that biological aging processes may increase optic nerve susceptibility to glaucomatous damage.

The characterization of fast and slow progressors in our study was rigorous and designed to create a clearly divergent sample, enhancing the ability to test our primary hypothesis. The marked contrast in rates of change between fast and slow progressors (−1.06 ± 1.08 dB/year vs −0.10 ± 0.33 dB/year in the fastest progressing eye, respectively) highlights the clinical significance of our findings and validates our sample construction strategy. Over the follow-up period, fast progressors lost an average of 8.2 dB in MD in their fastest progressing eye, compared to only 1.4 dB in the slow progressor group, highlighting the substantial difference in disease trajectory between these groups.

As expected, all epigenetic clocks demonstrated strong correlations with chronological age in our study population, with Pearson correlation coefficients ranging from 0.70 to 0.85. Following previously established methods, we calculated age acceleration as the residual from a linear regression model of each epigenetic clock and chronological age.^25^ This allowed us to quantify the degree to which an individual’s biological age, as measured by DNA methylation patterns, deviates from their chronological age. Using these age acceleration measures, our results demonstrated that individuals with faster glaucoma progression exhibited accelerated biological aging, as indicated by the various epigenetic clocks. The Horvath clock showed the strongest association, with fast progressors demonstrating a mean age acceleration of almost 3 years compared to slow progressors. This finding was further supported by the multivariable logistic regression model, which showed that for each year of age acceleration according to the Horvath clock, the odds of fast progression increased by 15% (OR: 1.15, 95% CI: 1.07-1.23, P<0.001), after controlling for other clinical and demographic variables that potentially affect progression speed.

The stronger association observed with the Horvath clock may be attributed to its “universal” nature. Horvath developed this epigenetic clock using samples from 51 different tissues and cell types, including brain, whole blood, heart, kidney, liver, lung, among others.^25^ While it did not specifically include eye tissue, it did incorporate brain tissue samples, which may be particularly relevant given the neurodegenerative nature of glaucoma. The Horvath clock’s ability to predict age across diverse tissues suggests it captures fundamental aging processes that may be applicable to the eye and optic nerve. This universality might explain its stronger association with glaucoma progression in our study, as it likely reflects systemic aging processes that could affect the vulnerability of retinal ganglion cells and their axons. The Hannum clock, specifically designed for blood samples, also showed a significant association, albeit weaker than the Horvath clock. Interestingly, the PhenoAge clock showed the weakest association, likely due to its construction. PhenoAge incorporates clinical phenotypes and mortality risk indicators, such as albumin, creatinine, glucose, and inflammatory markers, which may not be directly linked to glaucomatous damage.^27^ While these markers are relevant to overall health and aging, their connection to the specific mechanisms of glaucoma may be limited. The GrimAge clock, which showed an intermediate association, was developed to predict mortality and health span using DNA methylation-based biomarkers for plasma proteins and smoking history.^28^ Some of these factors, like smoking, may be relevant to glaucoma progression, while others may not directly influence optic nerve vulnerability, potentially explaining its intermediate performance.

The association between accelerated biological age and faster glaucoma progression may be explained by several mechanisms. Advanced biological age could increase the susceptibility of the optic nerve to damage due to a reduced ability to handle oxidative stress, which is recognized as a contributing factor to glaucomatous neurodegeneration.^1^ Aging is associated with mitochondrial dysfunction and decreased antioxidant defenses, potentially making retinal ganglion cells more vulnerable to IOP-related stress.^29–33^ Additionally, age-related biomechanical changes in the lamina cribrosa and peripapillary sclera could alter the optic nerve head response to IOP, potentially increasing susceptibility to axonal damage.^14, 34^

Our study found that age acceleration remained a significant predictor of fast glaucoma progression, even after adjusting for known risk factors such as IOP and baseline disease severity. The association of IOP and baseline MD with fast progression found in our models is similar to previous studies, supporting our approach. However, the independent association of epigenetic age acceleration suggests that biological aging contributes additional risk beyond these established factors. The observed differences in epigenetic age acceleration between fast and slow progressors were clinically significant, as depicted in the probability plot (**Figure 3**). This plot, based on the Horvath clock and adjusted for key confounders such as sex, race, peak IOP, baseline disease severity, CCT, and follow-up time, demonstrated a strong association between age acceleration and the likelihood of fast glaucoma progression. A 10-year increase in age acceleration was associated with a substantial increase in the probability of fast progression, nearly doubling it. For example, at an age acceleration of approximately +10 years, the probability of fast progression approached 70%. These findings highlight the potential clinical importance of epigenetic age acceleration as a predictor of glaucoma progression, beyond the influence of established risk factors.

**Figure 3.**
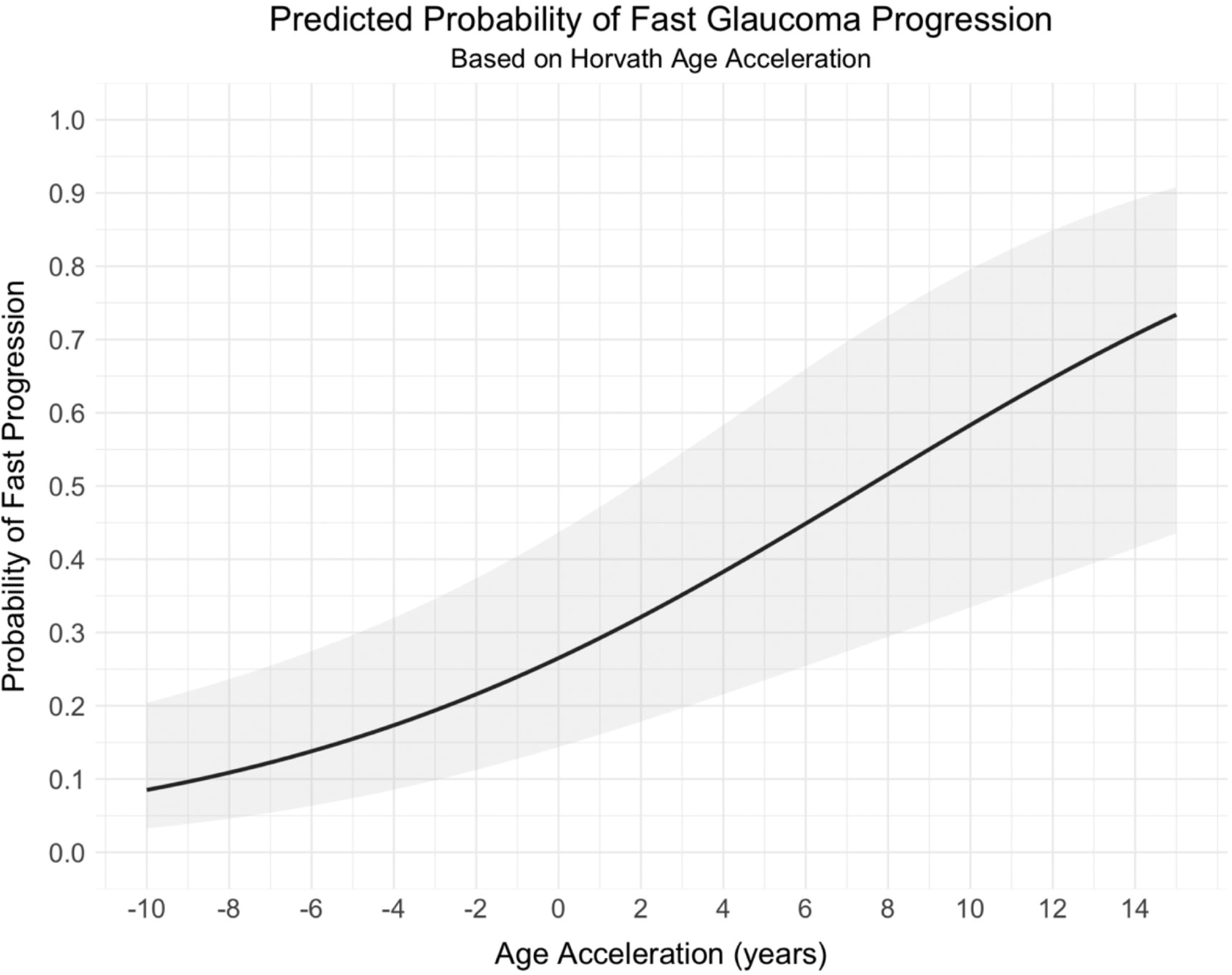
Predicted probability of fast glaucoma progression based on Horvath age acceleration The curve shows the predicted probability of fast glaucoma progression (y-axis) as a function of Horvath age acceleration (x-axis), derived from the multivariable logistic regression model. Age acceleration was calculated as the residual from a linear regression of Horvath epigenetic age on chronological age. The shaded area represents the 95% confidence interval. The model is adjusted for sex, race, peak intraocular pressure, baseline mean deviation, central corneal thickness and follow-up time until blood collection.

A particularly interesting finding was the stronger association between age acceleration and fast progression in subjects with low IOP, i.e., those who never had IOP greater than 21mmHg. This aligns with clinical observations that low-pressure glaucoma is rarely seen in young individuals and supports the notion that biological aging may increase the optic nerve susceptibility to damage even at normal IOP levels. The mean difference in AgeAccelHorvath between fast and slow progressors in this subgroup was almost 5 years, substantially larger than in the overall cohort, emphasizing the potential importance of biological aging in these cases.

The association between accelerated biological aging and increased susceptibility to damage is not unique to glaucoma and has been observed in other neurodegenerative diseases. For instance, in Alzheimer’s disease (AD), previous studies have found associations between epigenetic age acceleration and disease risk or progression.^35, 36^ Levine et al. reported that individuals with greater epigenetic age acceleration in brain tissue showed more rapid progression of AD neuropathology.^35^ Similarly, McCartney et al. found that DNA methylation age acceleration in blood was associated with cognitive decline and dementia risk.^36^ In Parkinson’s disease, another neurodegenerative condition, Horvath and Ritz reported that patients exhibited accelerated epigenetic aging in brain tissues compared to controls.^37^ These parallels with other neurodegenerative diseases support the notion that accelerated biological aging may increase susceptibility to neuronal damage across various conditions, including glaucoma.

The association between accelerated biological aging and glaucoma progression suggests potential for neuroprotective strategies targeting aging processes. Compounds like nicotinamide mononucleotide (NMN) and nicotinamide, precursors to NAD+, have shown promise in animal studies. Williams et al.^38^ demonstrated that nicotinamide administration prevented glaucoma in aged mice and those with ocular hypertension. Recent pilot human studies have also shown potential benefits. Hui et al.^39^ conducted a phase 2 clinical trial demonstrating that high-dose oral nicotinamide improved inner retinal function in glaucoma patients. Additionally, De Moraes et al.^40^ reported that oral nicotinamide supplementation enhanced retinal ganglion cell function in patients with early-stage glaucoma. While these results are promising, larger and longer-term studies are needed to fully establish the efficacy and safety of these compounds in human glaucoma patients. Nonetheless, these findings, coupled with our observations on epigenetic age acceleration, highlight the potential of targeting aging processes as a novel therapeutic approach in glaucoma management.

While our study provides compelling evidence for the role of biological aging in glaucoma progression, several limitations must be acknowledged. Primarily, blood samples for epigenetic analysis were collected after progression had occurred, precluding the establishment of a causal relationship. Although we attempted to control for this by including follow-up time to blood collection in our multivariable models, prospective studies are needed to confirm the predictive value of epigenetic age acceleration in glaucoma progression. Another limitation is the potential impact of glaucoma treatment on our results. While we controlled for IOP in our models, the effects of various treatments on epigenetic age acceleration are unknown and could potentially confound our findings. Additionally, our study was conducted in a single center, which may limit the generalizability of our results to other populations.

In conclusion, this study provides the first evidence of an association between accelerated epigenetic aging and faster glaucoma progression. Our findings suggest that biological age, as measured by DNA methylation patterns, may be an important factor in determining an increased susceptibility to glaucomatous damage. Future prospective studies are needed to validate these findings and explore the potential of epigenetic age as a biomarker for glaucoma progression.

## Data Availability

All data produced in the present study are available upon reasonable request to the authors.

## Abbreviations

AgeAccel: Age acceleration
CCT: central corneal thickness
CpG: cytosine-phosphate-guanine
DGR: Duke Glaucoma Registry
GAT: Goldmann applanation tonometry
IOP: intraocular pressure
ICD: international classification of diseases
MD: mean deviation
POAG: primary open-angle glaucoma
RNFL: retinal nerve fiber layer
SAP: standard automated perimetry
SITA: Swedish Interactive Thresholding Algorithm

## REFERENCES

1. Weinreb RN, Aung T, Medeiros FA. The pathophysiology and treatment of glaucoma: a review. JAMA 2014;311(18):1901–11.

2. Susanna BN, Ogata NG, Jammal AA, et al. Corneal Biomechanics and Visual Field Progression in Eyes with Seemingly Well-Controlled Intraocular Pressure. Ophthalmology 2019;126(12):1640–6.

3. Jammal AA, Berchuck SI, Thompson AC, et al. The Effect of Age on Increasing Susceptibility to Retinal Nerve Fiber Layer Loss in Glaucoma. Invest Ophthalmol Vis Sci 2020;61(13):8.

4. Mitchell P, Smith W, Attebo K, Healey PR. Prevalence of open-angle glaucoma in Australia. The Blue Mountains Eye Study. Ophthalmology 1996;103(10):1661–9.

5. Varma R, Ying-Lai M, Francis BA, et al. Prevalence of open-angle glaucoma and ocular hypertension in Latinos: the Los Angeles Latino Eye Study. Ophthalmology 2004;111(8):1439–48.

6. Collaborative Normal-Tension Glaucoma Study Group. Comparison of glaucomatous progression between untreated patients with normal-tension glaucoma and patients with therapeutically reduced intraocular pressures. Am J Ophthalmol 1998;126(4):487–97.

7. Leske MC, Connell AM, Wu SY, et al. Distribution of intraocular pressure. The Barbados Eye Study. Arch Ophthalmol 1997;115(8):1051–7.

8. Rochtchina E, Mitchell P, Wang JJ. Relationship between age and intraocular pressure: the Blue Mountains Eye Study. Clin Exp Ophthalmol 2002;30(3):173–5.

9. Friberg TR, Lace JW. A comparison of the elastic properties of human choroid and sclera. Exp Eye Res 1988;47(3):429–36.

10. Quigley HA, Addicks EM. Regional differences in the structure of the lamina cribrosa and their relation to glaucomatous optic nerve damage. Arch Ophthalmol 1981;99(1):137–43.

11. Grossniklaus HE, Nickerson JM, Edelhauser HF, et al. Anatomic alterations in aging and age-related diseases of the eye. Invest Ophthalmol Vis Sci 2013;54(14):ORSF23-7.

12. Ramrattan RS, van der Schaft TL, Mooy CM, et al. Morphometric analysis of Bruch’s membrane, the choriocapillaris, and the choroid in aging. Invest Ophthalmol Vis Sci 1994;35(6):2857–64.

13. Flammer J, Orgul S, Costa VP, et al. The impact of ocular blood flow in glaucoma. Prog Retin Eye Res 2002;21(4):359–93.

14. Burgoyne CF, Downs JC, Bellezza AJ, et al. The optic nerve head as a biomechanical structure: a new paradigm for understanding the role of IOP-related stress and strain in the pathophysiology of glaucomatous optic nerve head damage. Prog Retin Eye Res 2005;24(1):39–73.

15. Gabelt BT, Kaufman PL. Changes in aqueous humor dynamics with age and glaucoma. Prog Retin Eye Res 2005;24(5):612–37.

16. Horvath S, Raj K. DNA methylation-based biomarkers and the epigenetic clock theory of ageing. Nat Rev Genet 2018;19(6):371–84.

17. Marioni RE, Shah S, McRae AF, et al. DNA methylation age of blood predicts all-cause mortality in later life. Genome Biol 2015;16(1):25.

18. Jammal AA, Berchuck SI, Mariottoni EB, et al. Blood Pressure and Glaucomatous Progression in a Large Clinical Population. Ophthalmology 2022;129(2):161–70.

19. Jammal AA, Thompson AC, Mariottoni EB, et al. Impact of Intraocular Pressure Control on Rates of Retinal Nerve Fiber Layer Loss in a Large Clinical Population. Ophthalmology 2021;128(1):48–57.

20. Jammal AA, Thompson AC, Mariottoni EB, et al. Rates of Glaucomatous Structural and Functional Change From a Large Clinical Population: The Duke Glaucoma Registry Study. Am J Ophthalmol 2021;222:238–47.

21. Youssif AA, Onyekaba NA, Naithani R, et al. Social history and glaucoma progression: the effect of body mass index, tobacco and alcohol consumption on the rates of structural change in patients with glaucoma. Br J Ophthalmol 2024.

22. Leite MT, Rao HL, Zangwill LM, et al. Comparison of the diagnostic accuracies of the Spectralis, Cirrus, and RTVue optical coherence tomography devices in glaucoma. Ophthalmology 2011;118(7):1334–9.

23. Aryee MJ, Jaffe AE, Corrada-Bravo H, et al. Minfi: a flexible and comprehensive Bioconductor package for the analysis of Infinium DNA methylation microarrays. Bioinformatics 2014;30(10):1363–9.

24. Pidsley R, CC YW, Volta M, et al. A data-driven approach to preprocessing Illumina 450K methylation array data. BMC Genomics 2013;14:293.

25. Horvath S. DNA methylation age of human tissues and cell types. Genome Biol 2013;14(10):R115.

26. Hannum G, Guinney J, Zhao L, et al. Genome-wide methylation profiles reveal quantitative views of human aging rates. Mol Cell 2013;49(2):359–67.

27. Levine ME, Lu AT, Quach A, et al. An epigenetic biomarker of aging for lifespan and healthspan. Aging (Albany NY) 2018;10(4):573–91.

28. Lu AT, Quach A, Wilson JG, et al. DNA methylation GrimAge strongly predicts lifespan and healthspan. Aging (Albany NY) 2019;11(2):303–27.

29. Lin MT, Beal MF. Mitochondrial dysfunction and oxidative stress in neurodegenerative diseases. Nature 2006;443(7113):787–95.

30. Tezel G. Oxidative stress in glaucomatous neurodegeneration: mechanisms and consequences. Prog Retin Eye Res 2006;25(5):490–513.

31. Crowston JG, Kong YX, Trounce IA, et al. An acute intraocular pressure challenge to assess retinal ganglion cell injury and recovery in the mouse. Exp Eye Res 2015;141:3–8.

32. Kong YX, van Bergen N, Bui BV, et al. Impact of aging and diet restriction on retinal function during and after acute intraocular pressure injury. Neurobiol Aging 2012;33(6):1126 e15–25.

33. Levkovitch-Verbin H, Vander S, Makarovsky D, Lavinsky F. Increase in retinal ganglion cells’ susceptibility to elevated intraocular pressure and impairment of their endogenous neuroprotective mechanism by age. Mol Vis 2013;19:2011–22.

34. Liu B, McNally S, Kilpatrick JI, et al. Aging and ocular tissue stiffness in glaucoma. Surv Ophthalmol 2018;63(1):56–74.

35. Levine ME, Lu AT, Bennett DA, Horvath S. Epigenetic age of the pre-frontal cortex is associated with neuritic plaques, amyloid load, and Alzheimer’s disease related cognitive functioning. Aging (Albany NY) 2015;7(12):1198–211.

36. McCartney DL, Stevenson AJ, Walker RM, et al. Investigating the relationship between DNA methylation age acceleration and risk factors for Alzheimer’s disease. Alzheimers Dement (Amst) 2018;10:429–37.

37. Horvath S, Ritz BR. Increased epigenetic age and granulocyte counts in the blood of Parkinson’s disease patients. Aging (Albany NY) 2015;7(12):1130–42.

38. Williams PA, Harder JM, Foxworth NE, et al. Vitamin B(3) modulates mitochondrial vulnerability and prevents glaucoma in aged mice. Science 2017;355(6326):756–60.

39. Hui F, Tang J, Williams PA, et al. Improvement in inner retinal function in glaucoma with nicotinamide (vitamin B3) supplementation: A crossover randomized clinical trial. Clin Exp Ophthalmol 2020;48(7):903–14.

40. De Moraes CG, John SWM, Williams PA, et al. Nicotinamide and Pyruvate for Neuroenhancement in Open-Angle Glaucoma: A Phase 2 Randomized Clinical Trial. JAMA Ophthalmol 2022;140(1):11–8.

