## Supplemental Table 4 for "Accelerated Epigenetic Aging is Associated with Faster Glaucoma Progression: A DNA Methylation Study"

**Table 4 (Suppl).** Epigenetic age acceleration differences between fast and slow glaucoma progressors without history of elevated intraocular pressure.

| **Age Acceleration (years)** | **Mean Difference** | **95% CI Lower** | **95% CI Upper** | **P-value** |
| --- | --- | --- | --- | --- |
| **Horvath** | 4.69 | 2.46 | 6.93 | <0.001 |
| **Hannum** | 2.13 | 0.31 | 3.95 | 0.022 |
| **PhenoAge** | 1.21 | -1.10 | 3.53 | 0.301 |
| **GrimAge** | 3.27 | 0.86 | 5.69 | 0.008 |

Note: Age acceleration measures correspond to residuals of the regression between epigenetic age and chronologic age at time of blood collection. Mean difference is calculated as (mean of fast progressors) - (mean of slow progressors). Positive values indicate that fast progressors had higher age acceleration than slow progressors. CI: confidence interval.
